# Scalable Markers for Early Cognitive Decline: Plasma p-tau217, Subjective Cognitive Concerns, and Digital Testing: Results from the A4/LEARN studies

**DOI:** 10.1101/2025.10.14.25338009

**Authors:** Babak Khorsand, Devin Teichrow, Elham Ghanbarian, Lukai Zheng, S. Ahmad Sajjadi, Crystal M Glover, Joshua D. Grill, Laura A. Rabin, Ali Ezzati

## Abstract

**Background and Objectives:** Although amyloid positron emission tomography (PET) and Cerebrospinal fluid (CSF) biomarkers remain the standard for confirming Alzheimer’s disease (AD) pathology, their use is impractical for screening or routine prognostic assessment. Plasma phosphorylated tau 217 (p-tau217), subjective cognitive concerns, and computerized cognitive testing are non-invasive, scalable, and feasible to implement in large populations. We tested whether these measures independently predict the onset of cognitive impairment and whether combining them improves prognostic accuracy.

**Methods:** We analyzed 1,071 cognitively unimpaired adults aged 65–85 years from the Anti-Amyloid Treatment in Asymptomatic Alzheimer’s Disease (A4) trial (amyloid-positive; solanezumab or placebo arms) and the parallel Longitudinal Evaluation of Amyloid Risk and Neurodegeneration (LEARN) cohort (amyloid-negative). At baseline, participants completed plasma p-tau217 measurement, the Cognitive Function Index (CFI), and the Cogstate Computerized Battery (CCB). Over 240 weeks of follow-up, incident impairment was defined as conversion from a Global Clinical Dementia Rating Score (CDR-GS) of 0 to 0.5 or higher. The predictive value of each measure for subsequent decline was examined after adjustment for demographic and genetic covariates.

**Results:** During the follow-up, 365 of 1,071 participants (34.1%) developed cognitive impairment. Higher plasma p-tau217 (per–standard-deviation increase) was associated with higher odds of converting to CDR-GS>0 across all cohorts: A4-Placebo (HR=1.56; 95% CI, 1.37-1.78), A4-Solanezumab (HR=1.46; 95% CI, 1.29-1.65), LEARN (HR=1.25; 95% CI, 1.05-1.48). Similarly, higher CFI predicted incident impairment: A4-Placebo (HR=1.59; 95% CI, 1.42-1.79), A4-Solanezumab (HR=1.67; 95% CI, 1.47-1.91), LEARN (HR=1.37; 95% CI, 1.12-1.68). Lower CCB also conferred higher risk: A4-Placebo (HR=0.76; 95% CI, 0.65-0.91), A4-Solanezumab (HR=0.73; 95% CI, 0.62-0.87), LEARN (HR=0.68; 95% CI, 0.53-0.87). In models including all three predictors, each remained independently associated with progression.

**Conclusion:** Plasma p-tau217, subjective cognitive concerns, and computerized cognitive testing each independently predicted progression to cognitive impairment in cognitively unimpaired older adults. Together, these non-invasive and scalable measures provide practical tools for risk stratification years before clinical diagnosis. Combining biological, subjective, and digital markers may support earlier detection in clinical care and enhance efficiency in prevention trial enrollment.

## Introduction

Alzheimer’s disease (AD) is a progressive neurodegenerative disorder characterized by a prolonged preclinical phase in which pathophysiologic processes such as amyloid-β (Aβ) accumulation and tau aggregation occur years before overt symptoms (1). Identifying individuals at greatest risk for near-term cognitive decline during this preclinical stage is critical for timely intervention, efficient trial designs, and, ultimately, identifying ideal candidates for disease-delaying treatments. Although amyloid positron emission tomography (PET) and Cerebrospinal fluid (CSF) biomarkers reliably detect these pathologies, their invasiveness, cost, and limited accessibility restrict their use for routine screening and risk stratification (2). Moreover, subjective and functional aspects of early disease manifestation are not captured by PET or CSF biomarkers. As a result, there is increasing interest in accessible, low-burden measures that could be deployed in primary care, community, and large-scale research settings. Prominent among these emerging alternatives are plasma biomarkers, particularly plasma levels of phosphorylated tau at threonine 217 (p-tau217), which has shown strong associations with AD pathology and clinical progression in multiple cohorts (3); subjective cognitive concerns, as measured by tools such as the Cognitive Function Index (CFI), which may capture subtle self-perceived decline before objective deficits are detectable on neuropsychological testing (4); and Computerized Cognitive Battery (CCB), which provides standardized, language-independent assessment, with growing evidence of sensitivity to early changes linked to amyloid and tau pathology (5). While each of these markers—biological, subjective, and digital—has individually been linked to risk of cognitive decline, the extent to which they contribute independent and complementary information within the same framework is not known. Establishing whether these measures provide complementary rather than redundant information in different populations is essential for building efficient, multidomain risk models. The Anti-Amyloid Treatment in Asymptomatic Alzheimer’s Disease (A4) Study, a large, multinational randomized controlled trial, and its parallel observational cohort, the Longitudinal Evaluation of Amyloid Risk and Neurodegeneration (LEARN) Study, provide a unique framework to address this gap. Together, these studies include a large, diverse sample of clinically unimpaired older adults with longitudinal follow-up, rigorous cognitive outcome assessment, and detailed biomarker characterization. In this study, we tested the hypothesis that plasma p-tau217, CFI, and CBB each independently predict progression from normal cognition to symptomatic impairment over five years, and that their combination yields superior predictive performance compared with any single measure. We also examined whether these associations differed by baseline Aβ status.

## Methods

### Study Design and Participants

We conducted a secondary analysis of data from a randomized clinical trial (A4) and a parallel observational cohort study (LEARN) (6). The A4 Study was a multicenter clinical trial conducted at 67 sites across the United States, Australia, Japan, and Canada. It aimed to evaluate whether solanezumab could slow cognitive decline in older adults with preclinical Alzheimer’s disease—defined as cognitively unimpaired individuals with elevated amyloid-beta (Aβ) levels on PET scans. The LEARN study enrolled individuals who were screened for A4 but did not meet the amyloid threshold for trial enrollment.

Full eligibility criteria for the A4 trial were previously reported (6). In brief, participants were between 65 and 85 years old, had Mini-Mental State Examination (MMSE) scores between 25 and 30, a Global Clinical Dementia Rating Score (CDR-GS) of 0, and Logical Memory II scores of 6–18 based on educational level. All participants were required to have a study partner and undergo Aβ PET imaging. Exclusion criteria included significant comorbid conditions, elevated suicide risk, and certain medication use.

Of 6,763 individuals who were prescreened, 4,486 (66%) underwent F-florbetapir PET imaging. A total of 1,209 (27%) individuals with high amyloid burden (Aβ positive) were enrolled in the A4 trial and randomized 1:1 to receive either intravenous solanezumab (n=578) or placebo (n=591). From the remainder of screened participants, 538 (12%) individuals with lower amyloid burden (Aβ negative) were enrolled in the LEARN study.

For this analysis, we selected participants with available Apolipoprotein E ε4 carrier (APOE4) status, baseline p-tau217 measurement, baseline CCB, baseline CFI, and final visit (Week 240) CDR-GS scores. The final analytic sample included 1,071 participants, including 386 in the A4 placebo group, 381 in the A4 solanezumab group, and 304 in the LEARN study. Figure 1 illustrates the flow of participants from initial screening through final analytic sample, including numbers excluded at each step and reasons for exclusion (e.g., missing biomarker or cognitive data.

**Figure 1.**
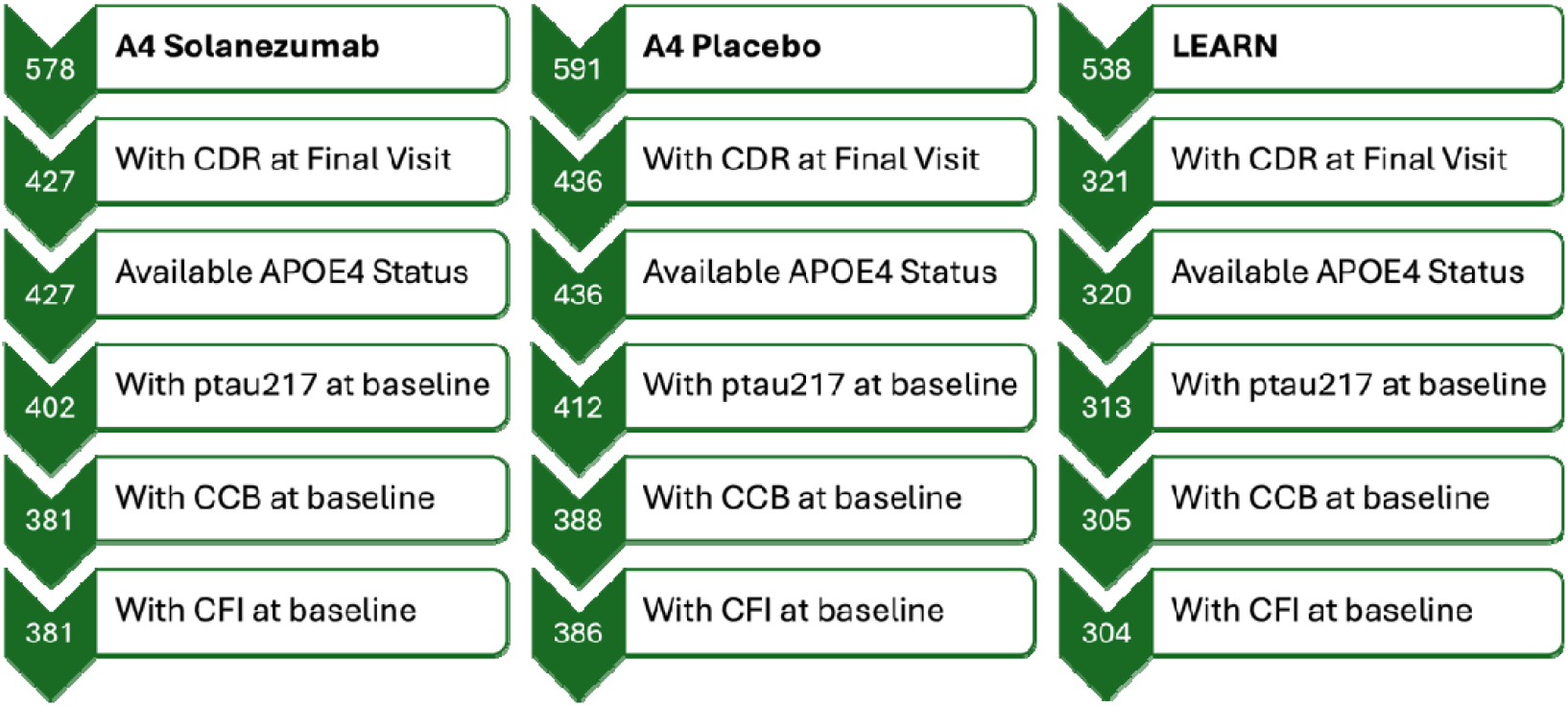
Flowchart of Study Participants. Abbreviations: CDR-GS = Clinical Dementia Rating Global Score; CCB = Cogstate Computerized Battery; CFI = Cognitive Function Index

### Study Measures

Baseline data were collected at enrollment (Week 0) for all measures, and cognitive status was assessed longitudinally, with final CDR-GS measured at Week 240.

#### Demographics

Demographic covariates included age, sex, and years of education. Age was analyzed as a continuous variable. Years of education was dichotomized as ≤12 years or >12 years.

#### Genetic Variable

APOE4 is the strongest genetic risk factor for Alzheimer’s disease. In the analysis, APOE4 status was included as a binary variable, with 0 representing individuals without the ε4 allele and 1 for those carrying at least one ε4 allele.

#### Imaging Biomarkers

Amyloid burden was quantified using 18F-florbetapir PET imaging. Quantitative measures were derived using the standardized uptake value ratio (SUVR), calculated by comparing tracer uptake in target cortical regions to a reference region. A threshold of SUVR>1.15, or SUVR between 1.1-1.15 with concordant visual read, was used to classify as amyloid-positive. Those below this threshold without visual read support were considered as amyloid-negative.

#### Plasma Biomarkers

p-tau217 levels were measured from plasma using an electro chemiluminescent immunoassay. Automated sample preparation was conducted using the Tecan Fluent workstation, and assays were run on the MSD Sector S Imager 600MM (7).

#### Cogstate Computerized Battery

The CCB is a set of standardized, computerized cognitive tests designed to assess various domains of cognitive function, including memory, attention, processing speed, and executive function (8). The composite score was derived from six tasks: the Behavioral Pattern Separation Object Test (recognition memory, higher scores indicating better performance); the Face Name Associative Memory Exam (associative memory, higher scores indicating better performance); the Detection Test (psychomotor speed, lower scores indicating better performance); the Identification Test (attention, lower scores indicating better performance); the One Card Learning Test (visual learning, higher scores indicating better performance); and the One Back Test (working memory, higher scores indicating better performance). For each task, raw scores were standardized to z-scores based on the study sample distribution, with directionality harmonized so that higher values consistently reflected better cognitive performance. The composite was then computed as the mean of these standardized scores.

Accordingly, the CCB composite should be interpreted as a continuous measure of global cognitive ability, where higher scores indicate better overall functioning.

#### Subjective Cognitive Concerns

Subjective cognitive concerns were assessed using the Cognitive Function Index (CFI), a modified version of the original 14-item questionnaire, which consists of 15 items assessing perceived cognitive decline in areas such as memory, language, and executive functioning) and functional abilities, relative to one year prior (9). Each item is scored as 0, 0.5, or 1, resulting in a total CFI score ranging from 0 to 15, with higher values indicating greater concern about cognitive decline.

#### Clinical Dementia Rating

The primary cognitive and functional endpoint was assessed using the CDR-GS. This instrument is based on semi-structured interviews conducted with both the participant and study partner, allowing evaluation of cognitive performance across six domains: memory, orientation, judgment and problem solving, community involvement, home and leisure activities, and personal care. Each domain is scored separately on a 5-point scale indicating impairment severity: 0 (none), 0.5 (very mild or questionable), 1 (mild), 2 (moderate), and 3 (severe dementia) [12]. For the present study, cognitive/functional decline was defined as a ≥0.5 increase in the CDR-GS during the 240-week follow-up period (10). Participants were followed for the CDR-GS test at weeks 48, 120, 168, 204, and 240.

### Statistical Analysis

Baseline characteristics were summarized descriptively. The primary outcome was incident symptomatic cognitive impairment, defined as progression from a baseline CDR-GS of 0 to a score of ≥0.5 at two consecutive follow-up visits or at the final visit (Week 240). Participants meeting this criterion were classified as Decliners; all others were considered Stable.

Survival analyses evaluated the association of plasma p-tau217, CFI, and CCB with risk of cognitive impairment. All models were adjusted for age, sex, years of education, and APOE ε4 status. Hazard ratios (HRs) with 95% confidence intervals (CIs) were reported. The proportional hazards assumption was tested with Schoenfeld residuals and was not violated. We estimated risk in each cohort (A4 Placebo, A4 Solanezumab, and LEARN) using a sequence of nested models: a base model with covariates only (age, sex, education, APOE ε4); base model plus each predictor separately (p-tau217, CFI, or CCB); base model with pairs of predictors; and a full model including all three predictors simultaneously. Each model estimated the hazard ratio (HR) and 95% confidence intervals (CI) for predictors of time to incident cognitive impairment. We modeled predictors in standard units, so all reported results represent hazard ratios per 1–standard deviation increase. Predictors were also modeled in their native units (p-tau217 as continuous assay values, CFI per 1-point increase, and CCB as composite score units), which are presented in Supplementary Table 1.

To visualize differences in time-to-event outcomes, we generated Kaplan–Meier survival curves, stratified by quartiles of p-tau217, CFI, and CCB. Group differences were assessed using the log-rank test.

## Results

### Sample Characteristics

The analytic sample included 1,071 participants aged 65 to 85 years at baseline (mean ± SD: 71.25 ± 4.5 years). On average, participants had 16.69 years of education (SD: 2.7). The cohort consisted of 59% women, and 49.9% of participants were APOE ε4 carriers. During the follow-up period of 240 weeks, 365 participants (34.1%) developed incident cognitive impairment, defined as an increase in CDR-GS from 0 at baseline to ≥0.5 at any follow-up. The remaining 706 participants (65.9%) remained cognitively stable.

In the entire sample, compared to stable participants, decliners were significantly older at baseline (72.7 vs 70.5, p<0.001), more likely to be male (51.5 vs 35.5, p<0.001), and more frequently APOE ε4 carriers (56.2% vs 46.6%, p = 0.03). They also had higher amyloid PET SUVR (1.33 vs 1.19, p<0.001), p-tau217 (0.31 vs 0.21, p<0.001), CFI (5.2 vs 2.7, p<0.001) and lower CCB (-0.16 vs 0.1, p<0.001). Sample characteristics, stratified by Stable and Decliners for each study, are summarized in Table 1.

**Table 1:** Baseline demographic, genetic, and biomarker characteristics of participants by cognitive outcome (Stable vs. Decliner) across the A4 Placebo, A4 Solanezumab, and LEARN cohorts.

|  | Solanezumab |  |  | Placebo |  |  | LEARN |  |  |
| --- | --- | --- | --- | --- | --- | --- | --- | --- | --- |
|  | Stable | Decliner | p | Stable | Decliner | p | Stable | Decliner | p |
| Samples, N (%) | 229 (60.1) | 152 (39.9) |  | 237 (61.4) | 149 (38.6) |  | 240 (78.9) | 64 (21.1) |  |
| Age, year (mean ± SD) | 70.88 ± 4.1 | 72.61 ± 4.8 | <0.001 | 70.76 ± 4.1 | 72.94 ± 5.2 | <0.001 | 69.85 ± 3.8 | 72.37 ± 5.3 | <0.001 |
| Sex, Male, N (%) | 89 (38.9) | 71 (46.7) | >0.05 | 80 (33.8) | 76 (51) | 0.019 | 82 (34.2) | 41 (64.1) | <0.001 |
| Education, year (mean ± SD) | 16.73 ± 2.5 | 16.52 ± 2.7 | >0.05 | 16.59 ± 2.9 | 16.56 ± 2.9 | >0.05 | 16.92 ± 2.5 | 16.78 ± 2.8 | >0.05 |
| BMI (mean ± SD) | 27.02 ± 4.9 | 27.47 ± 5.2 | >0.05 | 27.92 ± 5.1 | 27.29 ± 5.2 | >0.05 | 27.65 ± 5.1 | 28.21 ± 4.1 | >0.05 |
| APOE4 Carrier, N (%) | 135 (59) | 100 (65.8) | >0.05 | 146 (61.6) | 90 (60.4) | >0.05 | 48 (20) | 15 (23.4) | >0.05 |
| Amyloid PET SUVR (mean ± SD) | 1.29 ± 0.2 | 1.4 ± 0.2 | <0.001 | 1.3 ± 0.2 | 1.39 ± 0.2 | <0.001 | 0.99 ± 0.1 | 0.99 ± 0.1 | >0.05 |
| Plasma p-tau217 (mean ± SD) | 0.24 ± 0.1 | 0.35 ± 0.2 | <0.001 | 0.24 ± 0.1 | 0.32 ± 0.2 | <0.001 | 0.16 ± 0.1 | 0.18 ± 0.1 | <0.001 |
| CFI (mean ± SD) | 2.93 ± 2.6 | 5.78 ± 4.5 | <0.001 | 2.57 ± 2.6 | 5.1 ± 3.5 | <0.001 | 2.63 ± 2.6 | 4.28 ± 3.7 | <0.001 |
| CCB (mean ± SD) | 0.07 ± 0.5 | -0.18 ± 0.6 | <0.001 | 0.05 ± 0.5 | -0.18 ± 0.6 | <0.001 | 0.18 ± 0.5 | -0.07 ± 0.6 | <0.001 |

Abbreviations: BMI=Body Mass Index; SUVR=Standardized Uptake Value Ratio; CFI=Cognitive Function Index; CCB=Cogstate Computerized Battery

### Prediction of Incident Cognitive Impairment (CDR-GS ≥0.5)

Cox proportional hazards models were used to evaluate the association between baseline predictors and time to incident cognitive impairment, defined as an increase in CDR-GS from 0 to ≥0.5. Analyses were conducted separately for the A4 Placebo, A4 Solanezumab, and LEARN cohorts. All models were adjusted for age, sex, years of education, and APOE4 carrier status. As predictors were modeled in different units, hazard ratio magnitudes are not directly comparable and so per-standard-deviation hazard ratios were used.

Model 1, which included only demographic covariates and APOE4 status, identified age as a consistent and significant predictor across all three cohorts: A4 Placebo (HR=1.42; 95% CI, 1.21-1.67; P<0.001), A4 Solanezumab (HR=1.36; 95% CI, 1.16-1.59; P<0.001), and LEARN (HR=1.56; 95% CI, 1.25-1.95; P<0.001). Sex was also a significant predictor in A4 Placebo (HR=1.62; 95% CI, 1.17-2.25; P<0.001) and LEARN (HR=2.77; 95% CI, 1.63-4.71; P<0.001), with males showing greater risk for progression.

Models 2 through 4 tested the independent contribution of each candidate predictor—plasma p-tau217, CFI, and CCB—when added to the base model. Across all cohorts, higher baseline p-tau217 was significantly associated with increased risk of cognitive impairment: A4-Placebo (HR=1.56; 95% CI, 1.37-1.78), A4-Solanezumab (HR=1.46; 95% CI, 1.29-1.65), and LEARN (HR=1.25; 95% CI, 1.05-1.48).

Higher CFI scores (indicating more subjective cognitive concerns) were also strongly associated with greater risk: A4-Placebo (HR=1.59; 95% CI, 1.42-1.79), A4-Solanezumab (HR=1.67; 95% CI, 1.47-1.91), LEARN (HR=1.37; 95% CI, 1.12-1.68). Lower CCB scores were also associated with greater risk: A4-Placebo (HR=0.76; 95% CI, 0.65-0.91), A4-Solanezumab (HR=0.73; 95% CI, 0.62-0.87), LEARN (HR=0.68; 95% CI, 0.53-0.87).

Models 5 through 7 incorporated combinations of two candidate predictors, and Model 8 included all three (p-tau217, CFI, and CCB) simultaneously. The full model (Model 8) demonstrated that each of the three predictors remained independently and significantly associated with time to incident cognitive impairment in all three cohorts (Table 2).

**Table 2:** Cox Proportional Hazard Models Predicting Time to the Development of Incident Cognitive Impairment (CDR-GS ≥0.5), with predictors standardized per 1-SD increase.

|  | Model 1 | Model 2 | Model 3 | Model 4 | Model 5 | Model 6 | Model 7 | Model 8 |
| --- | --- | --- | --- | --- | --- | --- | --- | --- |
| **A4 Placebo** |  |  |  |  |  |  |  |  |
| Age | 1.42 ***<br>(1.21-1.67) | 1.32 ***<br>(1.12-1.55) | 1.34 ***<br>(1.15-1.57) | 1.31 **<br>(1.11-1.55) | 1.27 **<br>(1.09-1.48) | 1.22 *<br>(1.04-1.45) | 1.27 **<br>(1.08-1.49) | 1.21 *<br>(1.02-1.42) |
| Sex, Male | 1.62 **<br>(1.17-2.25) | 1.87 ***<br>(1.34-2.61) | 1.47 *<br>(1.06-2.06) | 1.61 **<br>(1.16-2.24) | 1.68 **<br>(1.19-2.36) | 1.88 ***<br>(1.34-2.62) | 1.45 *<br>(1.04-2.03) | 1.68 **<br>(1.19-2.36) |
| Education | 1.00<br>(0.95-1.06) | 1.00<br>(0.95-1.06) | 1.00<br>(0.95-1.07) | 1.01<br>(0.96-1.07) | 1.01<br>(0.95-1.06) | 1.01<br>(0.96-1.07) | 1.01<br>(0.96-1.08) | 1.01<br>(0.96-1.07) |
| APOE4 | 1.11<br>(0.80-1.55) | 1.06<br>(0.76-1.48) | 1.16<br>(0.83-1.63) | 1.12<br>(0.81-1.56) | 1.16<br>(0.82-1.61) | 1.08<br>(0.77-1.52) | 1.16<br>(0.83-1.62) | 1.17<br>(0.83-1.64) |
| p-tau217 |  | 1.56 ***<br>(1.37-1.78) |  |  | 1.57 ***<br>(1.38-1.79) | 1.54 ***<br>(1.35-1.76) |  | 1.56 ***<br>(1.37-1.78) |
| CFI |  |  | 1.59 ***<br>(1.42-1.79) |  | 1.62 ***<br>(1.43-1.83) |  | 1.56 ***<br>(1.39-1.76) | 1.59 ***<br>(1.41-1.79) |
| CCB |  |  |  | 0.76 **<br>(0.65-0.91) |  | 0.78 **<br>(0.66-0.92) | 0.82 *<br>(0.69-0.96) | 0.84 *<br>(0.71-0.99) |
| **A4 Solanezumab** |  |  |  |  |  |  |  |  |
| Age | 1.36 ***<br>(1.16-1.59) | 1.24 *<br>(1.05-1.46) | 1.33 ***<br>(1.14-1.56) | 1.23 *<br>(1.04-1.45) | 1.22 *<br>(1.03-1.44) | 1.16 *<br>(0.98-1.38) | 1.25 **<br>(1.06-1.48) | 1.17 *<br>(0.98-1.39) |
| Sex, Male | 1.19<br>(0.86-1.66) | 1.29<br>(0.93-1.79) | 0.98<br>(0.71-1.37) | 1.19<br>(0.85-1.65) | 1.07<br>(0.77-1.49) | 1.24<br>(0.89-1.73) | 1.01<br>(0.73-1.41) | 1.08<br>(0.78-1.50) |
| Education | 0.96<br>(0.91-1.03) | 0.97<br>(0.91-1.04) | 1.00<br>(0.94-1.06) | 0.99<br>(0.93-1.06) | 1.01<br>(0.94-1.07) | 0.99<br>(0.93-1.07) | 1.01<br>(0.95-1.08) | 1.01<br>(0.95-1.08) |
| APOE4 | 1.38<br>(0.98-1.94) | 1.25<br>(0.79-1.77) | 1.38<br>(0.98-1.93) | 1.39<br>(0.99-1.96) | 1.31<br>(0.93-1.84) | 1.27<br>(0.89-1.80) | 1.38<br>(0.98-1.93) | 1.31<br>(0.93-1.85) |
| p-tau217 |  | 1.46 ***<br>(1.29-1.65) |  |  | 1.36 ***<br>(1.19-1.55) | 1.41 ***<br>(1.24-1.59) |  | 1.34 ***<br>(1.18-1.52) |
| CFI |  |  | 1.67 ***<br>(1.47-1.91) |  | 1.60 ***<br>(1.40-1.83) |  | 1.63 ***<br>(1.42-1.86) | 1.57 ***<br>(1.37-1.80) |
| CCB |  |  |  | 0.73 ***<br>(0.62-0.87) |  | 0.78 **<br>(0.66-0.93) | 0.83 *<br>(0.70-0.99) | 0.86 *<br>(0.73-0.99) |
| **LEARN** |  |  |  |  |  |  |  |  |
| Age | 1.56 ***<br>(1.25-1.95) | 1.59 ***<br>(1.27-1.99) | 1.58 ***<br>(1.26-1.99) | 1.42 **<br>(1.12-1.80) | 1.60 ***<br>(1.27-2.01) | 1.45 **<br>(1.14-1.85) | 1.44 **<br>(1.13-1.84) | 1.46 **<br>(1.14-1.87) |
| Sex, Male | 2.77 ***<br>(1.63-4.71) | 2.68 ***<br>(1.58-4.54) | 2.37 **<br>(1.38-4.07) | 3.03 ***<br>(1.78-5.14) | 2.30 **<br>(1.34-3.96) | 2.91 ***<br>(1.72-4.95) | 2.59 ***<br>(1.51-4.47) | 2.52 ***<br>(1.46-4.34) |
| Education | 0.93<br>(0.84-1.03) | 0.93<br>(0.84-1.02) | 0.94<br>(0.86-1.04) | 0.94<br>(0.86-1.04) | 0.94<br>(0.86-1.04) | 0.94<br>(0.85-1.04) | 0.95<br>(0.87-1.05) | 0.95<br>(0.87-1.05) |
| APOE4 | 1.62<br>(0.87-3.02) | 1.49<br>(0.79-2.82) | 1.58<br>(0.85-2.93) | 1.58<br>(0.85-2.96) | 1.44<br>(0.76-2.71) | 1.50<br>(0.79-2.83) | 1.54<br>(0.83-2.87) | 1.43<br>(0.76-2.70) |
| p-tau217 |  | 1.25 *<br>(1.05-1.48) |  |  | 1.21 *<br>(1.01-1.45) | 1.21 *<br>(1.02-1.43) |  | 1.18 *<br>(1.00-1.41) |
| CFI |  |  | 1.37 **<br>(1.12-1.68) |  | 1.34 **<br>(1.09-1.65) |  | 1.32 **<br>(1.08-1.62) | 1.30 *<br>(1.06-1.59) |
| CCB |  |  |  | 0.68 **<br>(0.53-0.87) |  | 0.69 **<br>(0.54-0.89) | 0.70 **<br>(0.54-0.90) | 0.71 **<br>(0.54-0.92) |

**Note.** Hazard ratios (HRs) with 95% confidence intervals (CIs) are shown for each cohort. All models adjust for age, sex, education, and APOE $\epsilon 4$ carrier status. Predictors were standardized so HRs represent risk per 1-standard-deviation increase, allowing comparison across measures.
Abbreviations: CFI=Cognitive Function Index; CCB=Cogstate Computerized Battery
Significant Codes: \*\*\* p<0.001; \*\* p<0.01; \* p<0.05.

Kaplan–Meier survival curves, stratified by quartiles of baseline measures, revealed significant associations between each predictor and time to incident cognitive impairment, as assessed by the log-rank test (Figure 2). In the A4 Placebo group, higher baseline CFI scores were associated with increased risk of cognitive decline (χ^2^ = 78.1, P<0.001; Figure 2a). Similarly, higher plasma p-tau217 levels (χ^2^ = 31.1, P<0.001; Figure 2b) and lower CCB scores (χ^2^ = 15.1, p = 0.002; Figure 2c) were significantly associated with faster progression. Comparable results were observed in the Solanezumab arm of the A4 study, with significant associations for CFI (χ^2^ = 67.6, P<0.001), p-tau217 (χ^2^ = 45.7, P<0.001), and CCB (χ^2^ = 21.7, P<0.001). The LEARN cohort also showed consistent findings, with CFI (χ^2^ = 12.6, p = 0.006), p-tau217 (χ^2^ = 13.2, p = 0.004), and CCB (χ^2^ = 9.7, p = 0.02) each significantly predicting time to cognitive impairment.

**Figure 2:**
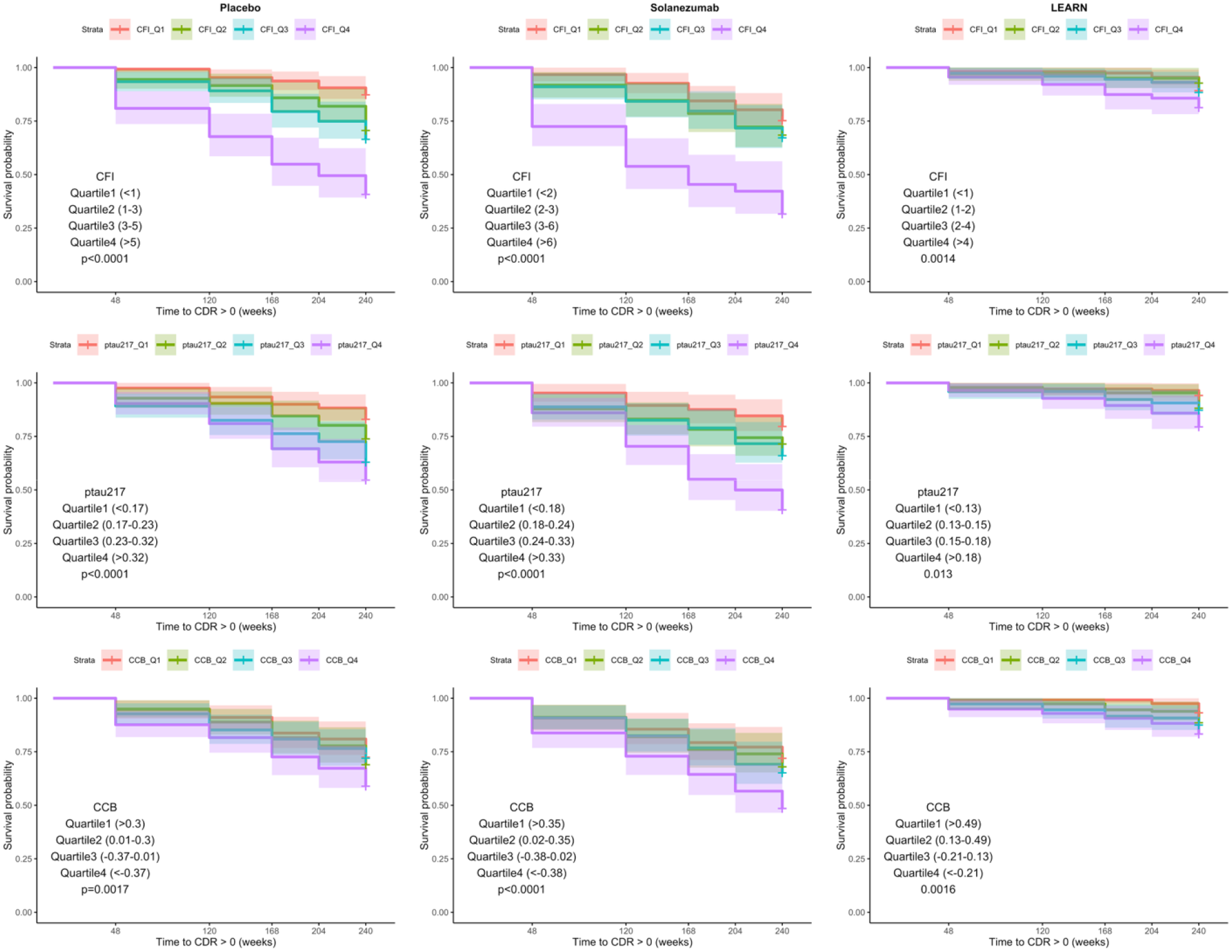
Kaplan–Meier curves for time to cognitive impairment (CDR-GS > 0) by quartiles of baseline measures. Notes: For CFI and plasma p-tau217, higher values indicate greater risk (risk increases Q1→Q4). For CCB, higher values indicate better cognition (risk increases Q4→Q1). Abbreviations: CFI=Cognitive Function Index; CCB=Cogstate Computerized Battery

## Discussion

In three well-characterized cohorts drawn from both a randomized clinical trial (A4) and a parallel observational study (LEARN), we found that plasma p-tau217, subjective cognitive concerns, and computerized cognitive testing each predicted progression to symptomatic impairment over nearly five years of follow-up. This multidomain framework—integrating biological, subjective, and digital measures— demonstrates that risk for near-term decline can be quantified using scalable tools accessible outside of specialized research settings. Importantly, predictive value was observed in both amyloid-positive and amyloid-negative participants, extending utility beyond biomarker-confirmed AD pathology. Differences in hazard ratio magnitudes across predictors reflect unit scaling rather than true differences in prognostic strength, and supplemental per-standard-deviation analyses confirmed that each measure contributed independent and complementary information. Together, these findings advance the field by showing that distinct, low-burden markers can be combined to improve risk stratification in preclinical populations.

While CSF and PET biomarkers have long been the gold-standard method for ante-mortem detection of AD neuropathologic change, their invasiveness, cost, and limited availability hinder widespread adoption. In contrast, p-tau217 offers a scalable, non-invasive alternative, measurable via simple blood draw on standard assay platforms. This accessibility enables early risk detection in primary care or community health settings lacking the gold-standard biomarker infrastructure. Multiple large-scale studies, including memory clinic cohorts (11) (12), diverse population samples, (13) and randomized trials (14) (15) have consistently demonstrated the strong diagnostic and prognostic utility of plasma p-tau217 across the AD continuum. Our findings add to this evidence: across two large multicenter cohorts with differing amyloid status, p-tau217 predicted clinical progression over nearly five years of follow-up and multiple CDR assessments. Its predictive power persisted when combined with subjective cognitive concerns and computerized testing, underscoring p-tau217 as a critical element in multidomain risk models.

The CFI questionnaire can be completed without a clinician, making it practical for large-scale screening. Despite its simplicity, the CFI reliably captures early cognitive changes: test-retest reliability is high, and rising CFI scores over time predict subsequent cognitive decline and worsening clinical dementia ratings (9, 16). These associations persist across diverse settings; cross-cultural validation studies demonstrate that the self-report version accurately identifies older adults with subjective cognitive decline and is easy to administer (17-19) while digital platforms show CFI scores correlate with daily functional impairments and higher risk of amyloid positivity (4). Given its low cost and strong predictive value, the CFI is well suited as a first-line tool to flag individuals who may benefit from more intensive cognitive testing or biomarker assessment—and our study confirms its predictive value by demonstrating that higher CFI scores were associated with clinical progression in our cohort.

Digital platforms are making cognitive assessment faster and more accessible for preclinical Alzheimer’s disease. Card-based tasks, self-rated memory tests, voice-driven questionnaires and even behavioral markers (gait, driving patterns, speech) can all be administered in minutes via tablets or smartphones and scored automatically. Evidence shows that a brief (<10 minutes) digital cognitive tests can separate cognitively unimpaired, amyloid-negative adults from amyloid-positive mild cognitive impairment and distinguish biomarker-negative mild cognitive impairment from prodromal AD, while self-rated memory and executive tests and voice-interaction screens each demonstrate high accuracy for detecting mild cognitive impairment and early dementia (20). Other studies also indicate that CCB capture early deficits, correlate with biomarker-defined risk and can be used for capturing cognitive decline in AD (21, 22). Our data show that a brief computerized cognitive test predicted progression to CDR-GS□≥ □0.5 over five years, and this association held across treatment, placebo and observational arms.

A key strength of this study is the simultaneous modelling of three accessible but distinct indicators of AD risk: plasma p-tau217, subjective cognitive concerns via the CFI, and computerized cognitive tests. Only a handful of studies have evaluated biological, subjective, and digital cognitive measures in the same framework; most examine these tests in isolation or in pairs. In one study, digital card-based tasks and remote memory tests correlated with plasma phosphorylated tau and combining them with p-tau181 yielded near-perfect discrimination between Alzheimer’s disease and control groups (23). Persistent subjective cognitive concerns amplify the dementia risk associated with glial fibrillary acidic protein, with odds ratios exceeding seven—substantially higher than those observed for either the biomarker or subjective complaints alone (24). Even after accounting for subjective cognitive decline, plasma p-tau217 and related blood markers remain significant predictors of preclinical cognitive deterioration in cognitively unimpaired elders (25). Together, these findings suggest that each domain of molecular pathology, digital biomarkers and subjective experience, captures unique aspects of preclinical Alzheimer’s disease.

Collectively, these findings suggest that molecular pathology, subjective assessment, and digital performance each capture unique aspects of the prodrome. Our study demonstrates that incorporating all three yields a more robust yet scalable risk model, providing a comprehensive picture of disease risk that can enhance early detection and intervention.

Blood-based prescreening can materially economize trial recruitment by reducing the number of negative amyloid PET scans; modelling suggests that triaging candidates with plasma p-tau assays can cut screen failures and associated costs by roughly 40–50% (26). Sequential strategies, such as using plasma p-tau217 to identify high-risk individuals and then confirming with tau PET—retain prognostic power while further improving efficiency(27). The predictors examined here, including plasma p-tau217, subjective cognitive concerns, and digital cognitive tests, are non-invasive, inexpensive and do not require specialist equipment. These characteristics make them attractive for broad adoption in primary care screening, digital cognitive health monitoring, population registries and decentralized clinical trials.

This work has several limitations. This study is a secondary analysis of A4/LEARN data with exploratory model choices; hence, findings should be considered hypothesis-generating rather than definitive. The highly educated, primarily non-Hispanic White volunteers in A4/LEARN limit generalizability to community populations and may understate access and other barriers to routine care. Missing data and attrition could bias estimates, and our outcome (conversion from CDR-GS 0 to ≥0.5) may miss more subtle preclinical changes. Lastly, each predictor has measurement constraints: the CBB can be affected by practice effects and device differences; the CFI could partially reflect mood or informant biases; and plasma p-tau217 assays vary across platforms. We did not examine longitudinal changes in these measures, so prospective validation is needed.

In summary, across the three A4/LEARN cohorts, baseline plasma p-tau217, subjective cognitive concerns, and a brief computerized cognitive battery each independently predicted conversion from CDR-GS 0 to ≥□0.5. Fully adjusted models showed that these markers offered complementary prognostic information, and survival curves separated consistently, indicating that inexpensive, scalable tools can flag elevated risk several years before clinical diagnosis. This supports a blood-first triage approach augmented by digital cognition and structured subjective report for early detection, counselling and trial enrichment. Future work should validate these findings in more diverse community cohorts, assess competing risks and missing-data mechanisms, examine device and platform effects, and determine whether longitudinal changes and composite risk scores can be integrated into primary-care workflows and decentralized trials.

## Data Availability

The data utilized in this article are from the publicly available dataset from the A4 trial, which can be accessed and downloaded from the Alzheimer's Clinical Trials Consortium (ACTC).

https://www.actcinfo.org/trial/a4-open-label-extension/

## Ethics declarations

All participants signed informed consent, and the current study does not meet the definition of human subject research.

## Author Contributions

Babak Khorsand (Conceptualization; Methodology; Writing - Original Draft; Formal analysis; Investigation); Devin Teichrow (Writing - Review & Editing); Elham Ghanbarian (Writing - Review & Editing), Lukai Zheng (Writing - Review & Editing); Joshua D. Grill (Writing - Review & Editing; Conceptualization); Crystal M. Glover (Writing - Review & Editing; Conceptualization); Seyed Ahmad Sajjadi (Writing - Review & Editing; Formal analysis); Laura Rabin (Writing - Review & Editing; Conceptualization); Ali Ezzati (Writing - Review & Editing; Formal analysis; Investigation; Conceptualization).

## Acknowledgements

This study was supported in part by grants from the National Institute of Health (NIA K23 AG063993; R01AG080635; R01AG095017); the Alzheimer’s Association (SG-24-988292 ISAVRAD); Cure Alzheimer’s Fund.

## Source of data and Funding

The A4 Study is a secondary prevention trial in preclinical Alzheimer’s disease, aiming to slow cognitive decline associated with brain amyloid accumulation in clinically normal older individuals. The A4 Study is funded by a public-private philanthropic partnership, including funding from the National Institutes of Health-National Institute on Aging (U19 AG010483, U24AG057437, R01 AG063689), Eli Lilly and Company, Alzheimer’s Association, Accelerating Medicines Partnership, GHR Foundation, an anonymous foundation and additional private donors, with in-kind support from Avid and Cogstate. The companion observational Longitudinal Evaluation of Amyloid Risk and Neurodegeneration (LEARN) Study is funded by the Alzheimer’s Association (LEARN-15-338729) and GHR Foundation. The A4 and LEARN Studies are led by Dr. Reisa Sperling at Brigham and Women’s Hospital, Harvard Medical School and Dr. Paul Aisen at the Alzheimer’s Therapeutic Research Institute (ATRI), University of Southern California. The A4 and LEARN Studies are coordinated by ATRI at the University of Southern California, and the data are made available through the Laboratory for Neuro Imaging at the University of Southern California. The participants screened for the A4 Study provided permission to share their de-identified data to advance the quest to find a successful treatment for Alzheimer’s disease.

---

**Supplementary Table 1.** Cox proportional hazards models predicting time to incident cognitive impairment (CDR-GS ≥ 0.5), with predictors modeled in their native measurement units.

|  | Model 1 | Model 2 | Model 3 | Model 4 | Model 5 | Model 6 | Model 7 | Model 8 |
| --- | --- | --- | --- | --- | --- | --- | --- | --- |
| **A4 Placebo** |  |  |  |  |  |  |  |  |
| **Age** | 1.08 ***<br>(1.04-1.12) | 1.06 ***<br>(1.03-1.09) | 1.07 ***<br>(1.03-1.10) | 1.06 **<br>(1.02-1.09) | 1.05 **<br>(1.02-1.09) | 1.04 *<br>(1.01-1.08) | 1.05 **<br>(1.02-1.09) | 1.04 *<br>(1.01-1.08) |
| **Sex, Male** | 1.62 **<br>(1.17-2.25) | 1.87 ***<br>(1.34-2.61) | 1.47 *<br>(1.06-2.06) | 1.61 **<br>(1.16-2.24) | 1.68 **<br>(1.19-2.36) | 1.88 ***<br>(1.34-2.62) | 1.45 *<br>(1.04-2.03) | 1.68 **<br>(1.19-2.36) |
| **Education** | 1.00<br>(0.95-1.06) | 1.00<br>(0.95-1.06) | 1.00<br>(0.95-1.07) | 1.01<br>(0.96-1.07) | 1.01<br>(0.95-1.06) | 1.01<br>(0.96-1.07) | 1.01<br>(0.96-1.08) | 1.01<br>(0.96-1.07) |
| **APOE4** | 1.11<br>(0.80-1.55) | 1.06<br>(0.76-1.48) | 1.16<br>(0.83-1.63) | 1.12<br>(0.81-1.56) | 1.16<br>(0.82-1.61) | 1.08<br>(0.77-1.52) | 1.16<br>(0.83-1.62) | 1.17<br>(0.83-1.64) |
| **p-tau217** |  | 24.71 ***<br>(9.55-63.94) |  |  | 26.79 ***<br>(10.49-68.41) | 22.63 ***<br>(8.69-58.95) |  | 25.09 ***<br>(9.75-64.61) |
| **CFI** |  |  | 1.16 ***<br>(1.11-1.19) |  | 1.16 ***<br>(1.12-1.21) |  | 1.15 ***<br>(1.11-1.19) | 1.15 ***<br>(1.11-1.20) |
| **CCB** |  |  |  | 0.60 **<br>(0.44-0.82) |  | 0.63 **<br>(0.46-0.86) | 0.68 *<br>(0.49-0.93) | 0.72 *<br>(0.52-0.98) |
| **A4 Solanezumab** |  |  |  |  |  |  |  |  |
| **Age** | 1.07 ***<br>(1.03-1.11) | 1.05 *<br>(1.01-1.09) | 1.07 ***<br>(1.03-1.10) | 1.05 *<br>(1.01-1.09) | 1.04 *<br>(1.01-1.08) | 1.03 *<br>(1.00-1.07) | 1.05 **<br>(1.01-1.09) | 1.04 *<br>(1.00-1.08) |
| **Sex, Male** | 1.19<br>(0.86-1.66) | 1.29<br>(0.93-1.79) | 0.98<br>(0.71-1.37) | 1.19<br>(0.85-1.65) | 1.07<br>(0.77-1.49) | 1.24<br>(0.89-1.73) | 1.01<br>(0.73-1.41) | 1.08<br>(0.78-1.50) |
| **Education** | 0.96<br>(0.91-1.03) | 0.97<br>(0.91-1.04) | 1.00<br>(0.94-1.06) | 0.99<br>(0.93-1.06) | 1.01<br>(0.94-1.07) | 0.99<br>(0.93-1.07) | 1.01<br>(0.95-1.08) | 1.01<br>(0.95-1.08) |
| **APOE4** | 1.38<br>(0.98-1.94) | 1.25<br>(0.79-1.77) | 1.38<br>(0.98-1.93) | 1.39<br>(0.99-1.96) | 1.31<br>(0.93-1.84) | 1.27<br>(0.89-1.80) | 1.38<br>(0.98-1.93) | 1.31<br>(0.93-1.85) |
| **p-tau217** |  | 9.81 ***<br>(4.69-20.46) |  |  | 6.38 ***<br>(2.93-13.91) | 7.81 ***<br>(3.74-16.31) |  | 5.81 ***<br>(2.68-12.59) |
| **CFI** |  |  | 1.15 ***<br>(1.11-1.19) |  | 1.13 ***<br>(1.09-1.17) |  | 1.14 ***<br>(1.10-1.18) | 1.13 ***<br>(1.09-1.17) |
| **CCB** |  |  |  | 0.55 ***<br>(0.40-0.76) |  | 0.63 **<br>(0.45-0.86) | 0.71 *<br>(0.51-0.97) | 0.75 *<br>(0.55-1.03) |
| **LEARN** |  |  |  |  |  |  |  |  |
| **Age** | 1.11 ***<br>(1.05-1.17) | 1.11 ***<br>(1.06-1.18) | 1.11 ***<br>(1.06-1.17) | 1.09 **<br>(1.03-1.15) | 1.12 ***<br>(1.06-1.18) | 1.09 **<br>(1.03-1.16) | 1.09 **<br>(1.03-1.15) | 1.09 **<br>(1.03-1.16) |
| **Sex, Male** | 2.77 ***<br>(1.63-4.71) | 2.68 ***<br>(1.58-4.54) | 2.37 **<br>(1.38-4.07) | 3.03 ***<br>(1.78-5.14) | 2.30 **<br>(1.34-3.96) | 2.91 ***<br>(1.72-4.95) | 2.59 ***<br>(1.51-4.47) | 2.52 ***<br>(1.46-4.34) |
| **Education** | 0.93<br>(0.84-1.03) | 0.93<br>(0.84-1.02) | 0.94<br>(0.86-1.04) | 0.94<br>(0.86-1.04) | 0.94<br>(0.86-1.04) | 0.94<br>(0.85-1.04) | 0.95<br>(0.87-1.05) | 0.95<br>(0.87-1.05) |
| **APOE4** | 1.62<br>(0.87-3.02) | 1.49<br>(0.79-2.82) | 1.58<br>(0.85-2.93) | 1.58<br>(0.85-2.96) | 1.44<br>(0.76-2.71) | 1.50<br>(0.79-2.83) | 1.54<br>(0.83-2.87) | 1.43<br>(0.76-2.70) |
| **p-tau217** |  | 39.91 *<br>(2.33-68.45) |  |  | 23.58 *<br>(1.16-48.12) | 22.89 *<br>(1.31-39.99) |  | 15.91 .<br>(0.80-31.81) |
| **CFI** |  |  | 1.11 **<br>(1.04-1.19) |  | 1.10 **<br>(1.03-1.18) |  | 1.10 **<br>(1.03-1.18) | 1.09 *<br>(1.02-1.17) |
| **CCB** |  |  |  | 0.46 **<br>(0.28-0.75) |  | 0.48 **<br>(0.29-0.79) | 0.49 **<br>(0.29-0.81) | 0.50 **<br>(0.30-0.84) |

Notes: Values are hazard ratios (HR) with 95% confidence intervals (CI).
Abbreviations: CFI=Cognitive Function Index; CCB=Cogstate Computerized Battery
Significant Codes: \*\*\* p<0.001; \*\* p<0.01; \* p<0.05.

